# SARS-CoV-2-Specific Antibody Profiles Distinguish Patients with Moderate from Severe COVID-19

**DOI:** 10.1101/2020.12.18.20248461

**Authors:** Leire de Campos Mata, Janet Piñero, Sonia Tejedor Vaquero, Roser Tachó-Piñot, Maria Kuksin, Itziar Arrieta Aldea, Natalia Rodrigo Melero, Carlo Carolis, Laura Furlong, Andrea Cerutti, Judit Villar-García, Giuliana Magri

## Abstract

The production of SARS-CoV-2-specific neutralizing antibodies is widely considered as a key mechanism for COVID-19 resolution and protection. However, beyond their protective function, antibodies to SARS-CoV-2 may also participate in disease pathogenesis. To explore the potential relationship between virus-specific humoral responses and COVID-19 immunopathology, we measured serum antibody classes and subclasses to the receptor-binding domain of the SARS-CoV-2 spike protein and the nucleoprotein in a cohort of hospitalized COVID-19 patients with moderate to severe disease. We found that RBD-specific IgG1 and IgG3 dominated the humoral response to SARS-CoV-2, were more abundant in severe patients, and positively correlated with several clinical parameters of inflammation. In contrast, a virus-specific IgA2 response skewed toward RBD rather than NP associated with a more favorable clinical course. Interestingly, RBD-dominant IgA2 responses were mostly detected in patients with gastrointestinal symptoms, suggesting the possible involvement of intrinsically tolerogenic gut immune pathways in the attenuation of virus-induced inflammation and disease resolution.

## Introduction

To date, the coronavirus disease 19 (COVID-19) pandemic has affected more than 70 million people, causing more than a million deaths worldwide (Glass et al., 2020). Following infection with SARS-CoV-2, the COVID-19 causative agent, individuals experience a wide variety of disease manifestations, ranging from asymptomatic state to severe disease with possible fatal outcomes (Guan et al., 2020; Wu and McGoogan, 2020; Zhou et al., 2020). Although the causes of such high diversity in disease severity are yet to be elucidated, increasing evidence suggests that, beyond genetic and demographic factors, the patient’s immune response to SARS-CoV-2 could play a central role in determining its clinical course. First of all, the innate and, especially, the adaptive immune responses are crucial for virus clearance and host protection (Poland et al., 2020; Tay et al., 2020). However, besides this beneficial function, host immune responses could also play a detrimental role in COVID-19 pathogenesis. Indeed, severe patients often display an acute respiratory distress syndrome, strongly associated with overaggressive inflammatory response and aberrant immune activation (Mehta et al., 2020; Tay et al., 2020).

Amid the different facets of the host immune response, virus-specific humoral immunity has been proposed to be critical in dictating the disease outcome. Virus-specific antibodies, directed against the SARS-CoV-2 trimeric spike (S) protein, which is required for virus entry, or the nucleocapsid protein (NP), which binds the viral RNA, emerge rapidly after infection (Long et al., 2020a). Early appearance of SARS-CoV-2-neutralizing antibodies, mainly directed against the receptor-binding domain (RBD) of the S protein has been reported in multiple studies (Piccoli et al., 2020; Robbiani et al., 2020; Suthar et al., 2020). Interestingly, the levels of SARS-CoV-2-specific antibodies and neutralizing titers are higher in individuals exhibiting more severe symptoms (Chen et al., 2020b; Guthmiller et al., 2020; Piccoli et al., 2020), raising the possibility that the quality of SARS-CoV-2-specific antibody response could influence not only the virus clearance but also the COVID-19 immunopathology. Indeed, beyond their protective function, antibodies could also have pathogenic extra-neutralizing properties that can lead to increased inflammation or antibody-dependent enhancement of infection (Iwasaki and Yang, 2020; Zohar and Alter, 2020). These extra-neutralizing functions strictly depend on the type of virus-specific antibody classes and subclasses induced upon infection (Zohar and Alter, 2020).

IgG antibodies consist of four subclasses termed IgG1, IgG2, IgG3, and IgG4. Each of these IgG subclasses has unique effector functions and properties that largely depend on their specific Fc domain and half-life. Virus-specific humoral responses are generally mediated by IgG1 and IgG3. These IgG subclasses are broadly considered pro-inflammatory, as they can initiate the complement cascade and deliver powerful immune-activating signals via Fcγ receptor I (FcγRI) and FcγRIII. Conversely, IgG2 and IgG4 subclasses are mostly non-inflammatory, as they neither activate complement nor signal through FcγRI and FcγRIII (Bournazos et al., 2020).

The characterization of both isotype and subclass profiles of the virus-specific antibody responses induced by the infection could also provide indirect information on the involvement of mucosal immunity. Among antibody isotypes, IgA is the most abundant class at mucosal surfaces (Chen et al., 2020a). In humans, IgA encompasses two distinct IgA1 and IgA2 subclasses. While IgA1 induction occurs in both mucosal and systemic compartments, IgA2 production is largely confined to intestinal segments (Chen et al., 2020a). Thus, detection of SARS-CoV-2-specific IgA2 could indicate the involvement of intestinal mucosal immune responses.

To explore the relationship between the SARS-CoV-2-specific antibody profile and COVID-19 immunopathology, we characterized serum antibodies to RBD of the viral spike protein and the NP in a cohort of hospitalized COVID-19 patients with moderate to severe disease. We found that RBD-specific IgG1 and IgG3 responses dominated humoral immunity in patients with more severe disease. In contrast, an IgA2 response skewed toward RBD rather than NP correlated with a more favorable clinical course. Interestingly, an RBD-dominant IgA2 response was mostly detected in patients with gastrointestinal symptoms, suggesting a possible involvement of gut immunity in disease resolution.

## Material and Methods

### Contact for Reagent and Resource Sharing

Further information and requests for resources and reagents should be directed to and will be fulfilled by the Lead Contact, Giuliana Magri. The dataset generated during and/or analyzed during the current study have been made available in the supplemental material.

### Experimental Model and Subject Details

#### Study Cohort

Serum samples from COVID-19 patients (N=38) were collected between March and May 2020 from the Mar Biobanc biorepository, affiliated to the Hospital del Mar (Barcelona, Spain), with patient informed consent. Only patients with confirmed SARS-CoV-2 infection by reverse transcription quantitative polymerase chain reaction (RT-qPCR) of nasopharyngeal swab were included. All sera were collected within the first 44 days following symptom onset, being the median time from symptom onset to blood testing 21 days [8-44]. The median age of patients was 61 [26-89] years old. 55 % were males. The median length of stay from hospitalization to discharge was 18 days [3-83]. Disease severity was determined according to the NIH Ordinal Severity score (range from 1 to 8) and Pulmonary affection severity score (range from 1 to 7), as previously described (Mathew et al., 2020). Clinical laboratory data were collected from the date closest to the first day of hospitalization. Demographic and clinical data of patients included in the study are summarized in **Table 1**. Sera from healthy controls (HCs) (N=12) collected prior to SARS-CoV-2 spread were also included in the study. All procedures followed were approved by the Ethical Committee for Clinical Investigation of the Institut Hospital del Mar d’Investigacions Mèdiques (CEIm núm 2020/9189/I).

#### Production of recombinant SARS-CoV-2 proteins

The pCAGGS RBD construct, encoding for the receptor-binding domain of the SARS-CoV-2 Spike protein (amino acids 319-541 of the Spike protein) along with the signal peptide plus a hexahistidine tag was provided by Dr Krammer (Mount Sinai School of Medicine, NY USA). The pLVX-EF1alpha-nCoV2019-N-2xStrep-IRES-Puro construct, encoding for the full-length SARS-CoV-2 nucleocapsid protein (NP) fused to a double Strep-tag at the C-terminus was a gift from Dr Krogan (University of California, San Francisco USA). Recombinant proteins were expressed in-house in Expi293F human cells (Thermo Fisher Scientific) by transfection of the cells with purified DNA and polyethylenimine (PEI). For secreted RBD proteins, cells were harvested 3 days post transfection and RBD-containing supernatants were collected by centrifugation at 13000rpm for 15min. RBD proteins were purified in Hitrap-ni Columns in an automated Fast Protein Liquid Chromatography (FPLC; Äkta avant), concentrated through 10 kDa Amicon centrifugal filter units (EMD Millipore) and resuspended in phosphate buffered saline (PBS). For NP, cell lysates from transfected cells were centrifuged at 13000 rpm for 20 min and the supernatant was loaded into a 5 ml Strep-Tactin XT column. The eluted nucleocapsid was concentrated through a 10 kDa Amicon centrifugal filter unit and purified with SEC (Sephadex 10/300) in PBS.

#### Enzyme-linked immunosorbent assay (ELISA)

ELISAs performed in this study were adapted from previously established protocols (Magri et al., 2017). Serum samples were heat-inactivated at 56ºC for 1 hour and stored at −80ºC prior to use. 96-well half-area flat bottom high-bind microplates (Corning) were coated overnight at 4ºC with each respective recombinant viral protein at 2ug/ml in PBS (30 ul per well) or with PBS alone. Plates were washed with PBS 0.05% Tween 20 (PBS-T) and blocked with blocking buffer (PBS containing 1.5% Bovine serum albumin, BSA) for 2 hours at room temperature (RT). Serum samples were serially diluted in PBS supplemented with 0.05% Tween 20 and 1% BSA and added to the viral protein-or PBS-coated plates for 2 hours at RT. After washing, plates were incubated with horseradish peroxidase (HRP)-conjugated anti-human Ig secondary antibodies diluted in PBS containing 0.05% Tween 20 1% BSA for 45 minutes at RT. Plates were washed 5 times with PBS-T and developed with TMB substrate reagent set (BD bioscience) with development reaction stopped with 1M H_2_SO_4_. Absorbance was measured at 450nm on a microplate reader (Infinite 200 PRO, Tecan). To detect RBD-specific and NP-specific IgM, IgA and IgG, HRP-conjugated anti-human IgA, IgG and IgM (Southern Biotech) were used at a 1:4000 dilution. To assess the distribution of the different IgG antibody subclasses, HRP-conjugated anti-human IgG1, IgG2, IgG3 and IgG4 (Southern Biotech) were used at a 1:3000 dilution. To detect SARS-CoV-2-specific IgA1 and IgA2, HRP-conjugated anti-human IgA1 or IgA2 (Southern Biotech) were used at a dilution of 1:2000 and 1:4000, respectively.

To quantitate the level of each viral antigen-specific antibody class or subclasses optical density (OD) values were calculated after subtraction of background (OD450 of serum dilutions on PBS-coated plates) and the area under the curve (AUC) was determined using Prism 8 (GraphPad). Negative threshold values were set using healthy control AUC levels plus 2 times the standard deviations of the mean.

### Data analysis and visualization

Analyses were performed using R (version 3.6.0, <u>R Core Team (2019)</u>. *<u>R: A Language and Environment for Statistical Computing</u>*<u>. Vienna, Austria: R Foundation for Statistical Computing</u>; https://www.R-project.org/.) and GraphPad Prism (version 8.0). Pairwise correlations between variables (including discrete ordinal, continuous scale, or a mixture of the two) were quantified by the Spearman rank correlation coefficient and visualized as a correlogram using R package corrplot (<u>Taiyun Wei and Viliam Simko (2017). R package “corrplot”: Visualization of a Correlation Matrix (Version 0.84)</u>). We display the positive correlations in red and negative correlations in blue. Spearman p-value significance levels were corrected using Benjamini-Hochberg method. The heatmaps were created using R package ComplexHeatmap (10.1093/bioinformatics/btw313), and row and column clustering were performed with complete cluster method and euclidean distance metric.

Recursive feature elimination using partial least square discriminant analysis (PLS-DA) from the Classification and Regression Training (<u>caret, Kuhn, M. caret: Classification and Regression Training. Astrophys. Source Code Libr. ascl:1505.003 (2015))</u> package in R was first used to select the best clinical and immunological predictors. We performed fivefold cross-validation during the training to provide robust and stable estimates. Two classification models were trained to distinguish moderate and severe disease groups using clinical variables and antibody titers as features, and antibody titers alone. We used the mixOmics R package to perform the visualizations related to the models (Rohart et al., 2017).

## Results

To investigate whether SARS-CoV-2 antibody responses differ among COVID-19 patients and elucidate how these responses relate to disease severity, we collected serum samples from a cohort of 38 hospitalized SARS-CoV-2-infected individuals with moderate to severe COVID-19. Sera from healthy controls (HCs) (N=12) collected prior to the beginning of SARS-CoV-2 spread were also included. All sera from infected individuals were collected between March and May 2020 at Hospital del Mar, Barcelona (Spain), within the first 44 days following symptom onset, being 21 days the median time from symptom onset to blood sampling [8-44]. A summary of demographic and clinical data of all patients included in this study is provided (**Table 1**).

To characterize specific profiles of antibody responses to SARS-CoV-2, we performed isotyping (IgA, IgG and IgM) and subtyping (IgG1, IgG2, IgG3, IgG4, IgA1 and IgA2) enzyme-linked immunosorbent assay (ELISA) using the receptor-binding domain (RBD) of the S protein as well as the nucleocapsid protein (NP) from SARS-CoV-2 as substrates. To quantify serum antibody concentrations, we performed six serial dilutions of serum samples and used optical density (OD) values to determine the area under the curve (AUC) (**Figure S1-2**).

First, we determined antibody responses to RBD, a domain of the viral S protein that strongly correlates with the neutralizing activity of virus-specific antibodies (Iyer et al., 2020; Ju et al., 2020; Suthar et al., 2020). Higher titers of both RBD-specific classes and subclasses were detected in COVID-19 patients compared to pre-pandemic HCs (**Figure S1**). Of note, less than 10% of patients showed RBD-specific IgG, IgA or IgM below threshold values. In a majority of patients, IgG1 and IgG3 subclasses dominated the RBD-specific IgG response. And indeed only 29% and 55% of patients showed RBD-specific IgG2 and IgG4 subclasses above threshold values, respectively. Remarkably, the RBD-specific IgA response encompassed both IgA1 and IgA2 subclasses in most patients. However, more patients induced RBD-specific IgA1 (92%) compared to RBD-specific IgA2 (79%).

Next, we dissected antibody responses to NP, a phosphoprotein from the viral nucleocapsid highly conserved among coronaviruses (**Figure S2**). Significantly higher titers of NP-specific IgM, IgG and IgA were detected in patients compared to HCs. Of note, NP-specific IgM was below threshold values in more patients (39%) compared to RBD-specific IgM (11%). In contrast, NP-specific IgG1 and IgG3 were detected in a majority of patients (> 92%), which reflected the pattern of RBD-specific IgG1 and IgG3. Unlike NP-specific IgG1 and IgG3, NP-specific IgG2 showed serum titers above the threshold value only in 68% of patients. Yet, NP-specific IgG2 was clearly present in a higher proportion of patients compared to HCs. Of note, NP-specific IgG4 titers were not significantly higher in patients compared to pre-pandemic HCs. Finally, the NP-specific IgA response included NP-specific IgA1 and, to a lesser degree, NP-specific IgA2.

To elucidate how distinct antibody profiles related to specific clinical outcomes, we performed hierarchical clustering of SARS-CoV-2 infected patients based on serum antibody titers to RBD and NP. This analysis was integrated with clinical and demographic data from each patient, such as sex, age, disease severity, hospitalization time, gastrointestinal symptoms and/or pulmonary fibrosis. Moreover, to establish whether specific antibody patterns could discriminate distinct infection stages, we included the time elapsed from symptom onset to blood sampling. This strategy allowed us to distinguish early, intermediate and late infection stages (**Figure 1A**).

**Figure 1.**
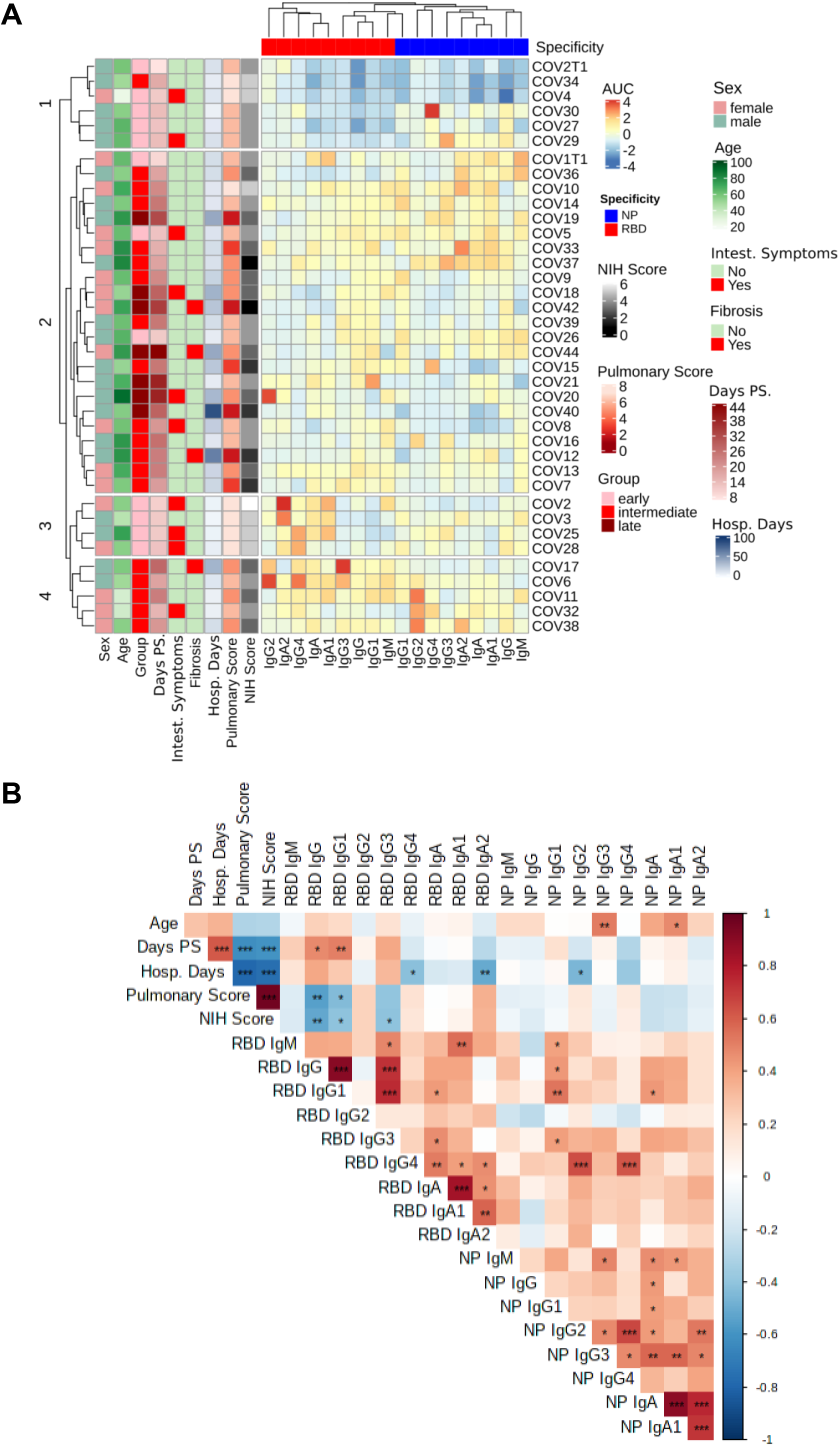
SARS-CoV-2-specific antibody profiles of COVID-19 patients relate with clinical variables. (**A**) Heat-map with the normalized values of SARS-CoV-2 RBD-specific and NP-specific Ig AUC (Z scored by column) of SARS-CoV-2 infected patients (n=38), clustered with hierarchical clustering. (**B**) Spearman’s correlation mapping between SARS-CoV-2-specific antibodies and clinical variables, including days of hospitalization, days post-symptoms (Days PS), Pulmonary Affection Severity score, and disease severity according to the NIH Ordinal Severity score (NIH Score). Spearman’s rank correlation coefficient (ρ) was indicated by heat scale; Spearman p-value significance levels were corrected using Benjamini-Hochberg method significance (*P < 0.05, **P < 0.01, and ***P < 0.001).

First, we analyzed how different virus-specific antibody isotypes and subclasses related to one another. Unsupervised hierarchical clustering indicated that RBD-specific IgA antibodies, including IgA1 and IgA2 subclasses, clustered together with non-inflammatory RBD-specific IgG2 and IgG4 subclasses but away from NP-specific antibodies, which instead clustered together with RBD-specific IgG1, IgG3 and IgM (**Figure 1A**).

With respect to the magnitude and properties of antibody responses to SARS-CoV-2, we identified four clusters of COVID-19 patients. Cluster 3 included patients at an early stage of the infection, who developed a dominant RBD-specific IgA response but low RBD-specific IgG1 and IgG3 responses. This antibody profile associated with a very favorable clinical outcome. Cluster 1, which also comprised patients with moderate disease, displayed low or absent RBD-specific IgG1 and IgG3 and a variable degree of NP-specific antibody responses. Cluster 2 was comprised of patients with more severe disease that mounted sustained RBD-specific IgG responses along with high or low NP-specific responses. Similar to Cluster 2, Cluster 4 included patients with more severe infection, but with sustained RBD-specific and NP-specific IgG responses that developed in parallel with virus-specific IgA responses (**Figure 1A**).

To further clarify associations between specific clinical outcomes and distinct antibody profiles we first performed integrated Spearman’s correlation mapping (**Figure 1B**). As expected, we found that RBD-specific total IgG titers positively correlated with RBD-specific IgG1 and IgG3 titers. In addition, RBD-specific IgG1 showed a robust positive correlation with RBD-specific IgG3 and with NP-specific IgG1. Remarkably, RBD-specific IgM and NP-specific IgM positively correlated with RBD-specific and NP-specific IgA1, suggesting coordinated IgA1 and IgM responses to SARS-CoV-2 at an early phase of the infection (**Figure 1B** and **Figure S3**). Despite its low titer, RBD-specific IgG4 was found to positively correlate with NP-specific IgG2 and IgG4. As described earlier, some patients also showed IgA2 responses to both RBD and NP viral proteins. Interestingly, RBD-specific IgA2 strongly correlated with RBD-specific IgA1 and to a lesser extent with RBD-specific IgG4, whereas NP-specific IgA2 correlated with NP-specific IgG2 in addition to NP-specific IgA1 (**Figure 1B** and **Figure S3)**.

Next, we investigated the relationship of distinct virus-specific antibody patterns with specific clinical and demographic properties of infected patients (**Figure 1B** and **S4**). We found that NP-specific IgA1 and IgG3 titers positively correlated with the age of patients **(Figure 1B** and **S4A)**. We also found that, unlike any other SARS-CoV-2-specific antibody, RBD-specific total IgG and RBD-specific IgG1 titers positively correlated with the time elapsed from symptom onset to blood sampling (**Figure 1B** and **S4B)**. To define disease severity, we considered the number of days of hospitalization along with two different clinical scores. The NIH Ordinal Severity score ranges from 1, which indicates a fatal outcome, to 8, which indicates no hospitalization and no limitation to physical activities. The Pulmonary Affection Severity score ranges from 1, which indicates severe acute respiratory distress syndrome and need of extracorporeal membrane oxygenation (ECMO), to 7, which indicates absence of any oxygen supplementation (**Table 1**). We found that the hospitalization time negatively correlated with RBD-specific IgA2 and, to a lesser extent, with RBD-specific IgG4 and NP-specific IgG2 (**Figure 1B** and **S4C)**. On the other hand, the NIH Ordinal Severity and Pulmonary Affection Severity scores negatively correlated with RBD-specific IgG, including RBD-specific IgG1 and IgG3 (**Figure S4 D-E)**.

To further explore possible associations between virus-specific antibody patterns and the disease severity, we considered various laboratory parameters indicative of inflammation extracted from the time closest to the first day of hospitalization. These parameters included the serum concentration of interleukin-6 (IL-6), ferritin, D-dimer, lactate dehydrogenase (LDH), C reactive protein (CRP) and blood neutrophil/lymphocyte ratio (**Table 1**). As reported by a published study (Mathew et al., 2020), all these laboratory parameters of inflammation but CRP positively correlated with different COVID-19 severity metrics (**Figure S5A**). Interestingly, RBD-specific IgG1 and IgG3 titers positively correlated with serum LDH (**Figure S5B-C)**. In addition, RBD-specific IgG3 titers showed a positive correlation with serum ferritin, whereas RBD-specific total IgG and IgG1 positively correlated with the neutrophil/lymphocyte ratio. Conversely, RBD-specific IgG4 negatively correlated with serum D-dimer. Interestingly, RBD-specific IgA2 titers inversely correlated with all clinical parameters of inflammation analyzed, although this correlation did not reach statistical significance after correction for multiple comparisons. No significant association was identified between clinical data and NP-specific antibody titers (**Figure S5B-C**).

Finally, we ran a machine learning latent variable modeling approach to define the minimal set of variables that could segregate individuals with different disease severity. First, we segregated COVID-19 patients into moderate patients with ≥4 NIH Ordinal Severity score and ≥6 Pulmonary Affection Severity score and severe patients with <4 NIH Ordinal Severity score and <6 Pulmonary Affection Severity score (**Table 1**). Next, we performed partial least square discriminant analysis (PLS-DA) upon combining clinical and immunological parameters. In this analysis, we included the relative overall ratios of RBD-specific:NP-specific antibody isotypes or subclasses. Indeed, selectively biased humoral responses were previously reported to be critical in determining a specific disease trajectory (Atyeo et al., 2020). Before building the model, we performed recursive feature elimination to keep only the feature most relevant to classification. Our model was 82% accurate at discriminating disease severity based on an area under the curve of the receiver-operator characteristic (ROC) curve analysis (**Figure 2A-C**). The most discriminative variables in the first component of the model of the high severity disease model included RBD-specific IgG and IgG1 titers, D-dimer, ferritin and RBD specific NP specific IgG3 ratio and were selected as the only relevant components after model tuning (**Figure 2D**). Conversely, RBD-specific:NP-specific IgA2 and gastrointestinal symptoms were the most discriminative first component variables of the moderate severity disease model (**Figure 2D**). Of note, PLS-DA performed after excluding laboratory clinical data from the analysis (**Figure S6A-C**) showed comparable accuracy. According to this new model (**Figure S6D**), RBD-specific IgG, IgG1 and the ratio RBD-specific:NP-specific IgG3 were the top three correlative variables of high disease severity. Moreover, the model not including clinical laboratory data correlated RBD-specific:NP-specific IgA2, RBD-specific IgA2 and gastrointestinal symptoms with moderate disease severity. In agreement with the models, RBD-specific IgG, IgG1 and IgG3 titers were significantly higher in patients with severe disease compared to patients with moderate disease (**Figure 2E)**, whereas RBD-specific IgG1 titers were lower in patients that did experience gastrointestinal symptoms (**Figure 2F**). Interestingly, patients with moderate disease and those experiencing intestinal symptoms showed a skewed RBD-specific IgA2 response (**Figure 2E-F**).

**Figure 2.**
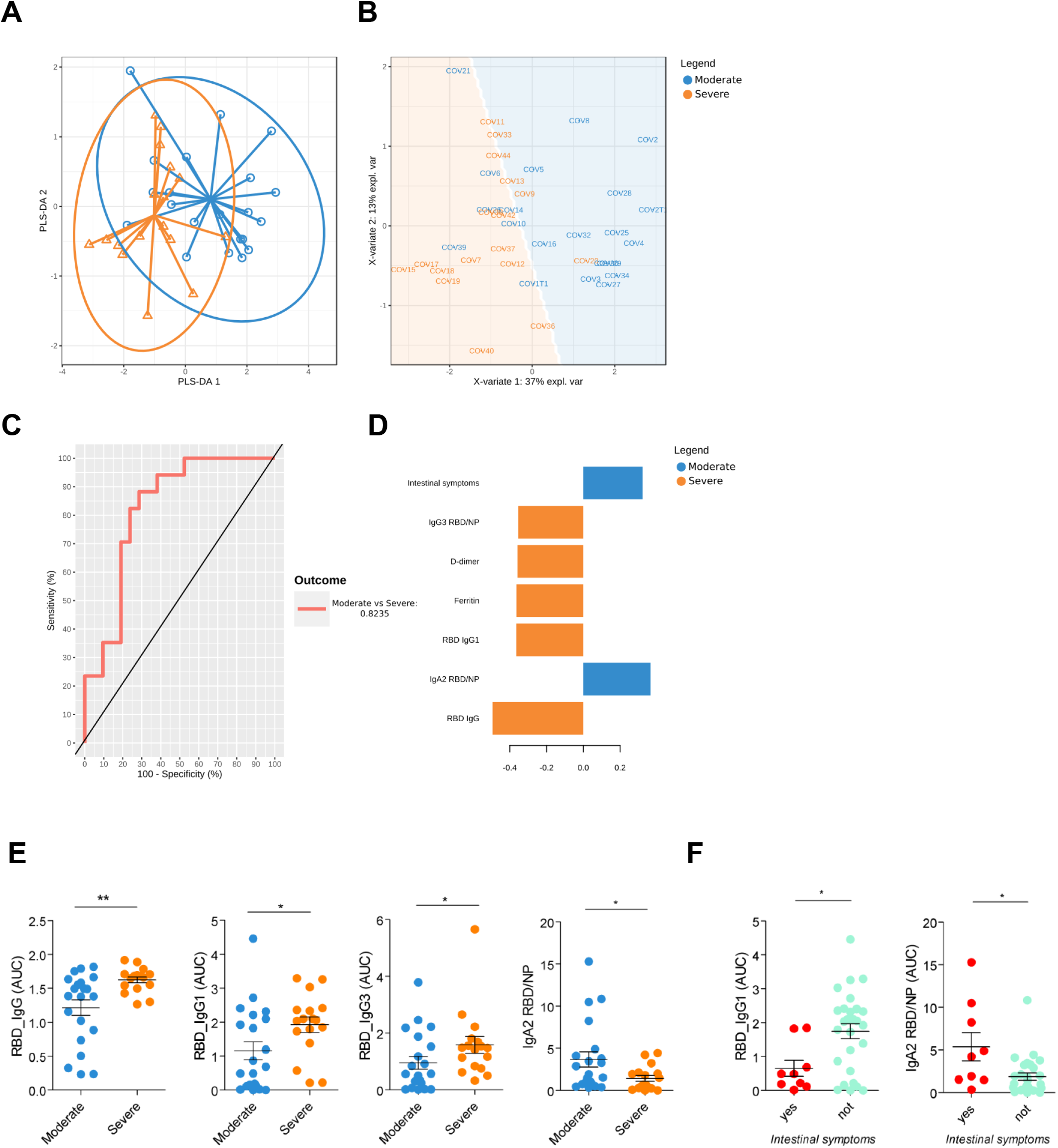
Partial least squares-discriminant analysis (PLS-DA) of SARS-CoV-2-specific antibody profiles and clinical variables distinguish patients with moderate from severe COVID-19. (**A**) Map of the patients into the first two dimensions of the PLS-DA model. The points represent dimension scores of the projection of high-dimensional feature vectors onto the first (x axis) and second (y axis) dimensions. Confidence ellipses for each class are plotted to highlight the strength of the discrimination (confidence level set to 95%). (**B**) Map of the patients on the first two dimensions of the PLS-DA model with the ‘prediction’ background. The algorithm estimates the predicted area for each class, defined as the 2D surface where all points are predicted to be of the same class. (**C**) ROC Curve of the classification performance of the PLS-DA model for the component one. The AUC is calculated from training cross-validation sets and averaged. (**D**) The loading weights of each variable on the first component of the PLS-DA model. The most important variables (according to the absolute value of their coefficients) are ordered from bottom to top. (**E**) SARS-CoV-2-specific Ig AUC values from serum samples of patients grouped based on disease severity (**F**) SARS-CoV-2-specific Ig AUC values from serum samples of patients grouped based on the presence or absence of intestinal symptoms. (**E**) and (**F**); two-tailed Mann-Whitney U test (*P < 0.05, **P < 0.01, and ***P < 0.001).

## Discussion

Our study provides a characterization with subclasses resolution of the antibody response against SARS-CoV-2 RBD and NP in a cohort of hospitalized COVID-19 patients with different disease severity. Here we show that IgG1 and IgG3 subclasses dominated the IgG response to SARS-CoV-2. As such, a larger magnitude of RBD-specific IgG1 and, to lower extent IgG3 titers, is linked to increased disease severity throughout the infection. Of note, our machine-learning model identified RBD-specific IgG titers as the most powerful parameters to discriminate disease severity among hospitalized patients, on top of other laboratory clinical data used to predict clinical outcomes, such as serum D-dimer and ferritin. In addition, we show that IgG2 and IgG4 subclasses represented a rather marginal component of the IgG response to SARS-CoV-2 and did not associate with disease severity or experience of pulmonary fibrosis, ruling out the possible involvement of IgG4-related lung disease in COVID-19 pathogenesis. Finally, we found that an RBD-dominant IgA2 response, mostly detected in patients with gastrointestinal symptoms, correlated with a more favorable clinical outcome.

It has been previously reported that the levels of SARS-CoV-2 specific antibodies and the neutralizing activity of sera are higher in hospitalized patients compared to SARS-CoV-2-infected asymptomatic individuals or COVID-19 patients experiencing mild symptoms (Chen et al., 2020b; Guthmiller et al., 2020; Piccoli et al., 2020). This difference in antibody titers has been mostly correlated with potential differences in immune protection. Our characterization of virus-specific antibody class and subclasses in COVID-19 hospitalized patients may provide some hints on the role of antibodies in COVID-19 immunopathology. Indeed, one possible explanation for the correlation of S-targeting IgG1 and IgG3 with disease severity may relate to their ability to exacerbate inflammation in the advanced phases of infection. Consistent with this possibility, we found that RBD-specific IgG1 and IgG3 positively correlated with canonical biomarkers of inflammation, such as serum LDH and ferritin as well as the neutrophil/lymphocyte ratio. Interestingly, COVID-19 patients with more severe disease have been recently shown to produce IgG antibodies with more prominent pro-inflammatory properties in their Fc domain, including afucosylated RBD-specific IgG1 and RBD-specific IgG3. When present in SARS-CoV-2-containing immunocomplexes, afucosylated IgG antibodies elicit production of pro-inflammatory cytokines by myeloid cells (Chakraborty et al., 2020). This effect involves a signaling pathway emanating from the FcγRIIIa and may cause enhanced pathology. While these are plausible explanations, RBD-specific IgG responses could merely reflect an increased antigenic burden as reported in other infections (Kawahara et al., 2019; Tomaras and Haynes, 2009).

In both SARS-CoV-1 and SARS-CoV-2 infection, humoral responses to the S protein arise in concomitance with antibody responses to NP (Atyeo et al., 2020; Guthmiller et al., 2020; Shi et al., 2004). However, despite some degree of correlation between RBD-specific IgG1 and NP-specific IgG1, NP-specific antibodies showed no correlation with disease severity. This observation is consistent with earlier reports (Atyeo et al., 2020; Guthmiller et al., 2020), but its biological underpinnings remain unclear.

In addition to virus-specific IgG, we found that SARS-CoV-2 infection induces the production of RBD-specific as well as NP-specific IgA and IgM antibodies. Of note, SARS-CoV-2-specific IgA1 and IgM responses highly correlate to one another, which may be due to the massive involvement of mucosal surfaces in the early phases of infection (Sterlin et al., 2020). However, as reported previously by others (Dogan et al., 2020; Zohar et al., 2020), we also found that virus-specific IgA1 and IgM showed no correlation with disease severity or any biomarkers of inflammation. Importantly, human IgA responses include IgA1 and IgA2 subclasses (Chen et al., 2020a). While IgA1 predominates in both intestinal and respiratory mucosae, IgA2 is largely confined to the intestinal mucosa (Chen et al., 2020a). Remarkably, a subset of COVID-19 patients showed IgA2 responses specific to RBD and NP in addition to virus-specific IgA1 responses, which may reflect the involvement of gut immune response, secondary to local infection. Gut infection by SARS-CoV-2 would be consistent with the elevated expression by intestinal epithelial cells of angiotensin-converting enzyme type 2 (ACE2), which is the host receptor for SARS-CoV-2 (Xiao et al., 2020; Zhang et al., 2020). Accordingly, recent studies have shown infection of intestinal enterocytes in COVID-19 patients (Livanos et al., 2020; Xiao et al., 2020). Moreover, SARS-CoV-2 genome and RBD-specific IgA was detected in the stool from COVID-19 patients, with more abundant viral RNA in patients suffering intestinal symptoms (Britton et al., 2020).

Recently, gastrointestinal symptoms presumably caused by intestinal SARS-CoV-2 infection were found to be associated with favorable clinical outcomes and increased survival rather than gut inflammation (Livanos et al., 2020). Consistent with this and other observations (Livanos et al., 2020; Wang et al., 2020), we found that RBD-specific IgA2 titers negatively correlated with the days of hospitalization. Moreover, skewed RBD-specific IgA2 responses increased in patients with a more favorable clinical outcome and intestinal symptoms. These results raise the possibility that gut immunity attenuates the pathogenicity of SARS-CoV-2 and promotes disease resolution. This conclusion needs to be validated by additional studies involving more patients. However, it could be speculated that early gut infection triggers an attenuated immune response to SARS-CoV-2 as a result of the intrinsically tolerogenic properties of the intestinal mucosa. Due to the extensive functional crosstalk between the intestinal and respiratory tracts, gut-driven immune responses to SARS-CoV-2 may lead to an attenuation of inflammation in the lungs of COVID-19 patients.

### Limitations of the study

There are a number of limitations in this study. First of all, it includes a limited number of COVID-19 patients (n=38) and presence of co-morbidities were not taken into consideration, thus our results should be confirmed in future studies in a larger cohort of SARS-CoV-2-infected patients. Our serologic analysis has been performed only on COVID-19 hospitalized patients ranging from moderate to severe COVID-19. Although previous reports (Guthmiller et al., 2020; Long et al., 2020b; Robbiani et al., 2020) have described striking differences in the magnitude of virus-specific humoral responses between asymptomatic and hospitalized individuals, mild patients may be analyzed in future studies to confirm the association between antibody profiles (i.e. RBD-specific IgA2) and intestinal symptoms or time of disease resolution. Finally, the time from symptom onset to blood sampling should be also considered as a possible confounder as for some patients with low virus-specific antibody titers, serologic analysis may have been performed before the subject had mounted a significant humoral response.

## Data Availability

De-identified raw data of the results of serological assays will be provided upon request.

## Author Contributions

L.D.C.M performed experiments, analyzed and discussed data and wrote the manuscript; J.P. performed data analysis, discussed data and wrote the manuscript; S.T.V. and R.T.P performed ELISA experiments and discussed data; M.K. performed initial data analysis; N.R.M. and C.C produced recombinant SARS-CoV-2 antigens; A.C. and L.F. discussed data and wrote the manuscript; J.V.G and I.A.A selected COVID-19 patients, provided clinical data and discussed data; G.M. designed and performed experiments, analyzed results, discussed data, and wrote the manuscript.

## Acknowledgments

We want to particularly acknowledge the patients and the Parc de Salut Mar MARBiobanc (PT17/0015/0011) integrated in the Spanish National Biobanks Network from ISCIII for their collaboration. MARBiobanc’s work was supported by grants from Instituto de Salud Carlos III/FEDER (PT17/0015/0011) and the “Xarxa de Bancs de tumors” sponsored by Pla Director d’Oncologia de Catalunya (XBTC). This study was supported by the COVID-19 call grant from Generalitat de Catalunya, Department of Health (to G.M and L.D.C.M) and grant Miguel Servet research program (to G.M). IMI-JU resources of which are composed of financial contribution from the EU-FP7 [FP7/2007–2013] and EFPIA companies in kind contribution [116030 to TransQST, 777365 to eTRANSAFE],and the EU H2020 Programme 2014–2020 [676559 to Elixir-Excelerate]; Agència de Gestió d’Ajuts Universitaris i de Recerca Generalitat de Catalunya [2017SGR00519]. The Research Programme on Biomedical Informatics (GRIB) is a member of the Spanish National Bioinformatics Institute (INB), funded by ISCIII and FEDER (PRB2-ISCIII [PT13/0001/0023, of the PE I+D+i 2013–2016]). The DCEXS is a ‘Unidad de Excelencia María de Maeztu’, funded by the MINECO [MDM-2014-0370].

## Notes

### Competing Interest Statement

The authors have declared no competing interest.

### Author Declarations

All procedures followed were approved by the Ethical Committee for Clinical Investigation of the Institut Hospital del Mar d Investigacions Mediques (CEIm number 2020/9189/I)

