## Supplemental materials for "SARS-CoV-2-Specific Antibody Profiles Distinguish Patients with Moderate from Severe COVID-19"

**Table 1**

**Supplemental Figures and Legends S1 – S6**

| VARIABLE | Moderate | Severe |
|---|---|---|
| Number of patients | 21 | 17 |
| **Demographics** | | |
| Age (yr) | 58 [26-73] | 65 [35-89] |
| Sex (F/M) | 8 (38%) / 13 (62%) | 9 (53%) / 8 (47%) |
| **Disease characteristics** | | |
| Number of days after symptom onset | 17 [8-38] | 26 [16-44] |
| Days of hospitalization | 9 [3-22] | 29 [5-83] |
| NIH Ordinal Severity Score | 4 [4-6] | 3 [1-3] |
| NIH Ordinal Severity Score | 4 [4: 14 (67%); 5: 6 (29%); 6: 1 (5%)] | 3 [1: 2 (12%); 2: 4 (24%); 3: 11 (65%)] |
| Pulmonary affection severity score | 6 [6-7] | 4 [2-5] |
| Pulmonary affection severity score | 6 [6: 14 (67%); 7: 7 (33%)] | 4 [2: 4 (24%); 3: 3 (18%); 4: 0 (0%); 5: 10 (59%)] |
| Fibrosis (y/n) | 0 (0%) / 21 (100%) | 4 (24%) / 13 (76%) |
| Intestinal symptoms (y/n) | 8 (38%) / 13 (62%) | 2 (12%) / 15 (88%) |
| **Biology** | | |
| D-dimer (µg/L FEU) | 1477 [310-7590] | 4589 [470-25610] |
| Ferritin (ng/mL) | 815 [172-2864] | 1558 [103-4873] |
| Lactate dehydrogenase (UI/L) | 277 [180-615] | 327 [175-500] |
| C reactive protein (mg/dL) | 6.8 [0.3-22.6] | 10.6 [0.3-32.3] |
| Interleukin 6 (pg/mL) | 103.9 [1.5-1279] | 532.8 [3.3-6462] |
| Neutrophil to lymphocyte ratio | 5.6 [0.9-20] | 8.5 [1.7-24.7] |

**Table 1. Demographic and clinical data of patients included in the study.** Median and range are shown for continuous variables in demographics, disease characteristics and laboratory clinical data. Count and proportion are shown for categorical variable modalities. NIH ordinal severity score for hospitalized patients ranges from not requiring supplemental oxygen-no longer requires ongoing medical care (6), not requiring supplemental oxygen-requiring ongoing medical care (5), requiring supplemental oxygen (4), on non-invasive ventilation or high flow oxygen devices (3), on invasive mechanical ventilation or extracorporeal membrane oxygenation (ECMO) (2) and death (1). Pulmonary affection severity score ranges from room air (RA) (7), nasal cannula (NC) (6), high flow nasal cannula – noninvasive ventilation (HFNC-NIV) (5), mild acute respiratory distress syndrome (ARDS) (4), moderate ARDS (3), severe ARDS (2), and severe ARDS with extracorporeal membrane oxygenation (ECMO)(1).

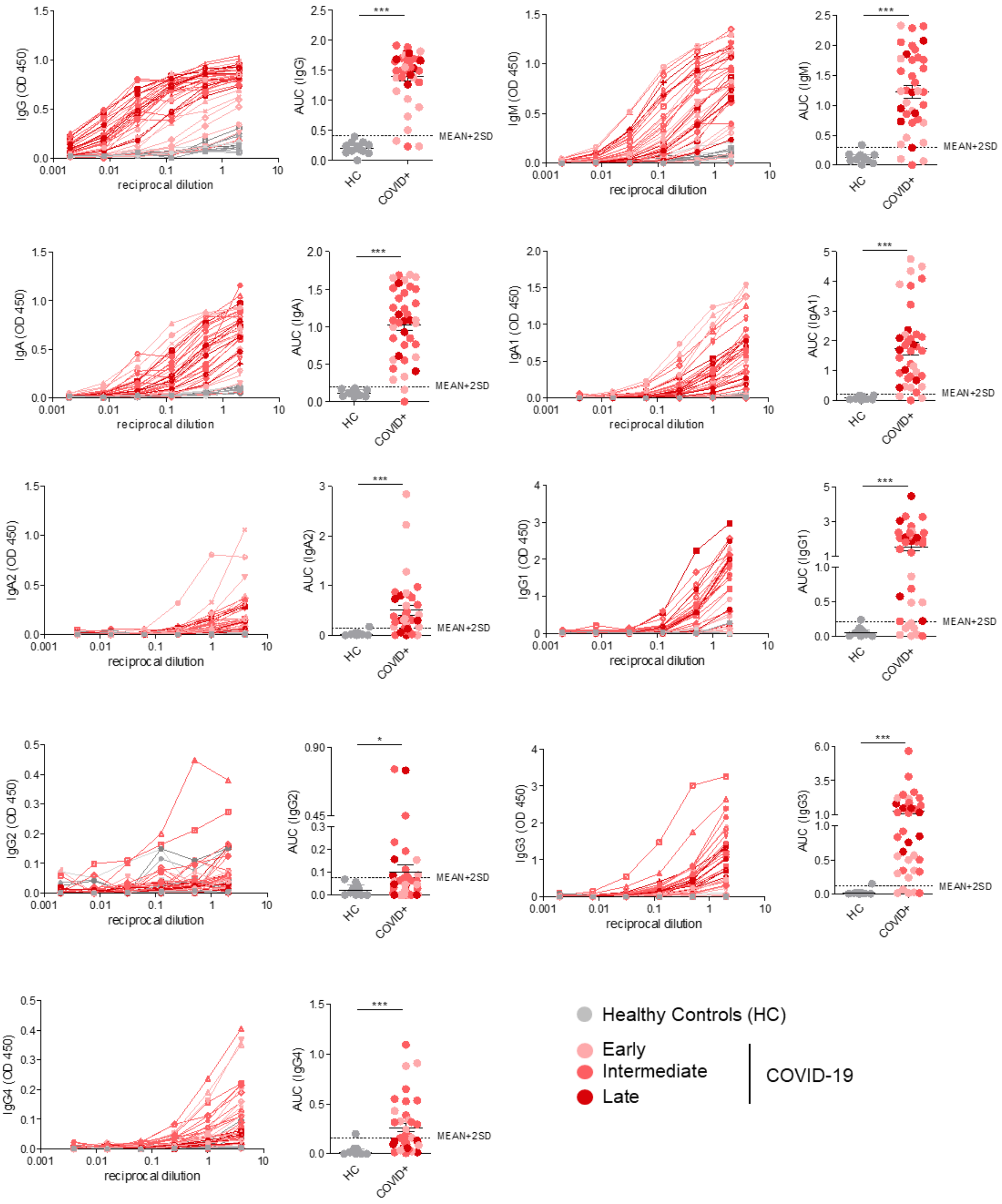


**Figure S1. SARS-CoV-2 RBD-specific humoral response.** For each of RBD-specific antibody classes and subclasses analyzed, the left panel depicts the titration of the individual serum samples and the right panel represents the area under the curve (AUC). Sera from healthy controls (HCs), collected prior to SARS-CoV-2 spread were analyzed in parallel to establish negative threshold values (healthy control AUC mean plus 2 times the standard deviations of the mean). Data are presented as mean ± SEM. Two-tailed Mann-Whitney U test (*P < 0.05, **P < 0.01, and ***P < 0.001).

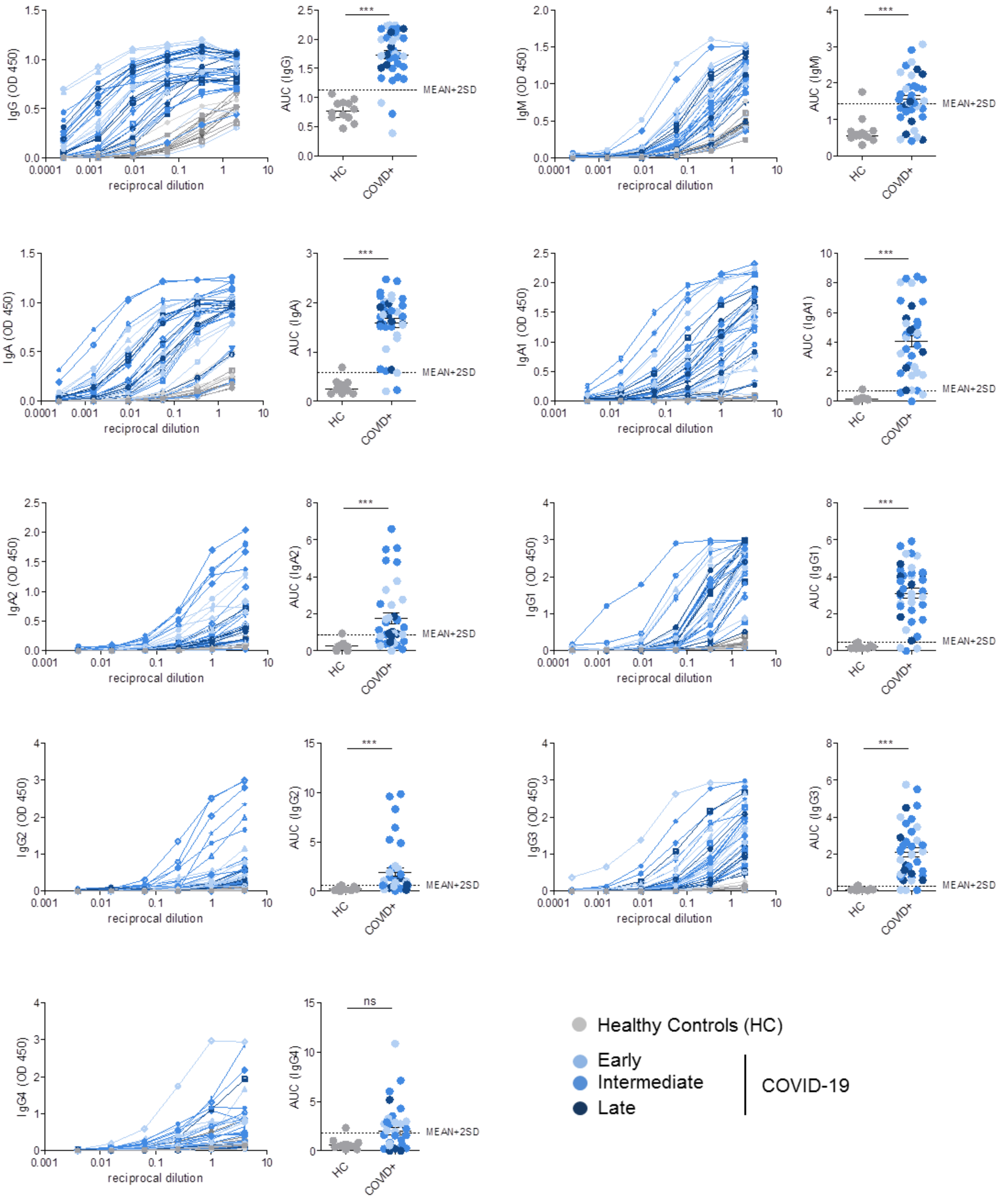


**Figure S2. SARS-CoV-2 NP-specific humoral response.** For each of the NP-specific antibody classes and subclasses analyzed, the left panel depicts the titration of the individual serum samples and the right panel represents the area under the curve (AUC) calculated from the same experiment. Sera from healthy controls (HCs), collected prior to SARS-CoV-2 spread were analyzed in parallel to establish negative threshold values (healthy control AUC mean plus 2 times the standard deviations of the mean). Data are presented as mean ± SEM. Two-tailed Mann-Whitney U test (*P < 0.05, **P < 0.01, and ***P < 0.001).

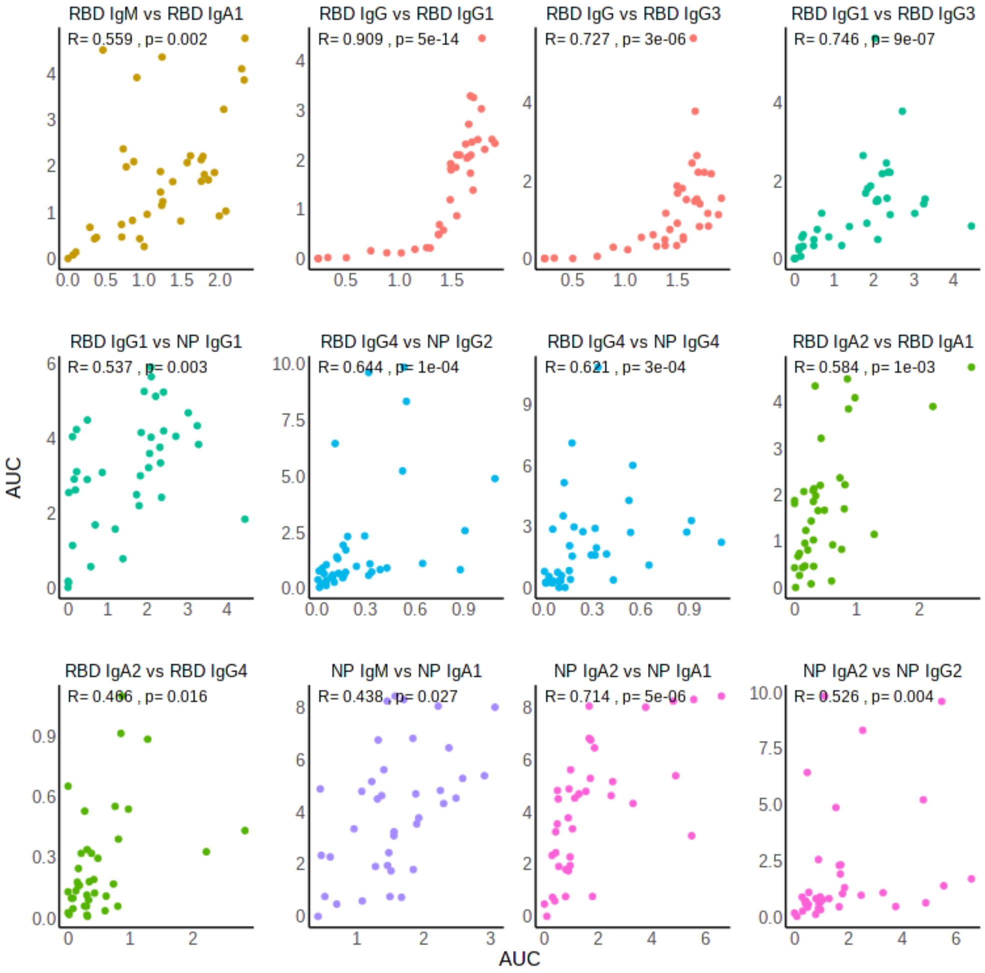


**Figure S3. Scatter plots depicting significant correlations between the AUC (area under the curve) of SARS-CoV-2-specific antibody classes and subclasses (FDR < 0.05) from COVID-19 patients.** Spearman correlation coefficients and the associated corrected p-values (Benjamini-Hochberg method) are shown.

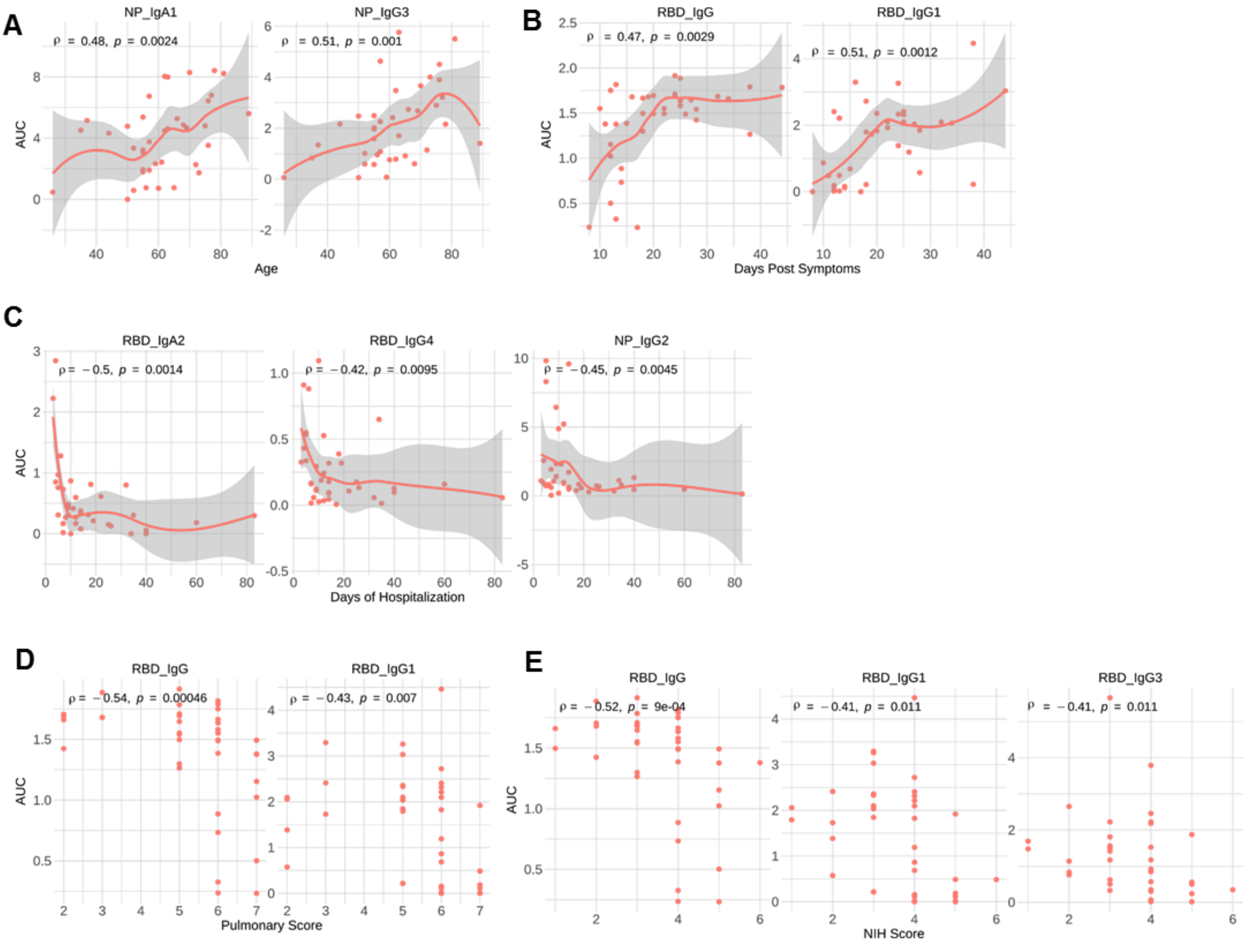


**Figure S4. Scatter plots depicting significant correlations between the AUC (area under the curve) of SARS-CoV-2-specific antibody classes and subclasses (FDR < 0.05) and clinical variables.** Correlations between serum concentration of the specified antibodies and (**A**) patient's age, (**B**) days post-symptoms, (**C**) days of hospitalization, (**D**) Pulmonary Affection score (range from 1 to 7) and (**E**) NIH Severity score (range from 1 to 6). Spearman correlation coefficients and the associated corrected p-values (Benjamini-Hochberg method) are shown.

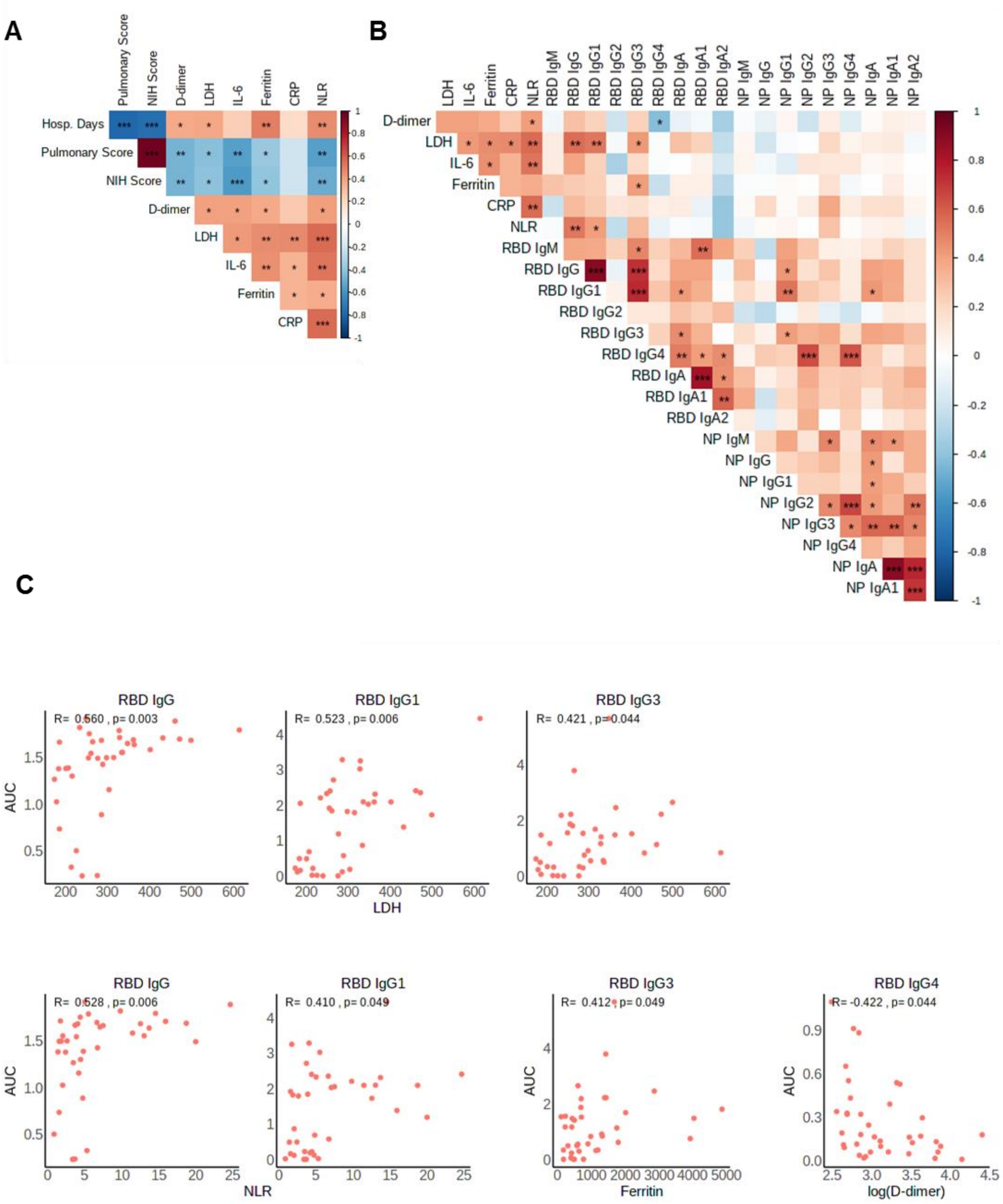


**Figure S5. Correlation between clinical variables, laboratory parameters indicative of inflammation and SARS-CoV-2-specific antibody classes and subclasses.** (**A**) Heatmap representing the correlation between the days of hospitalization (Hosp. Days), the disease scores (the NIH Ordinal Severity score (NIH Score) and the Pulmonary Affection Severity score (pulmonary score)) with the blood clinical variables: D-dimer, lactate dehydrogenase (LDH), IL-6, ferritin, C-reactive protein (CRP), and neutrophils/lymphocytes ratio (NLR). (**B**) Heatmap representing the correlation between the serum concentration of virus-specific immunoglobulin classes and subclasses with the blood clinical variables: D-dimer, Lactate dehydrogenase (LDH), IL-6, ferritin, C-reactive Protein (CRP), and neutrophils/lymphocytes ratio (NLR). Spearman's rank correlation coefficient (ρ) is indicated by heat scale, with red or blue color indicating a positive or negative correlation respectively. Corrected p-values (Benjamini-Hochberg method) is indicated by *P < 0.05, **P < 0.01, and ***P < 0.001. (**C**) Scatter plots depicting the significant correlations between the plasma concentration of virus specific Ig and blood clinical variables: LDH, NLR, ferritin and D-dimer. Spearman correlation coefficients and the associated corrected p-values (Benjamini-Hochberg method) are shown.

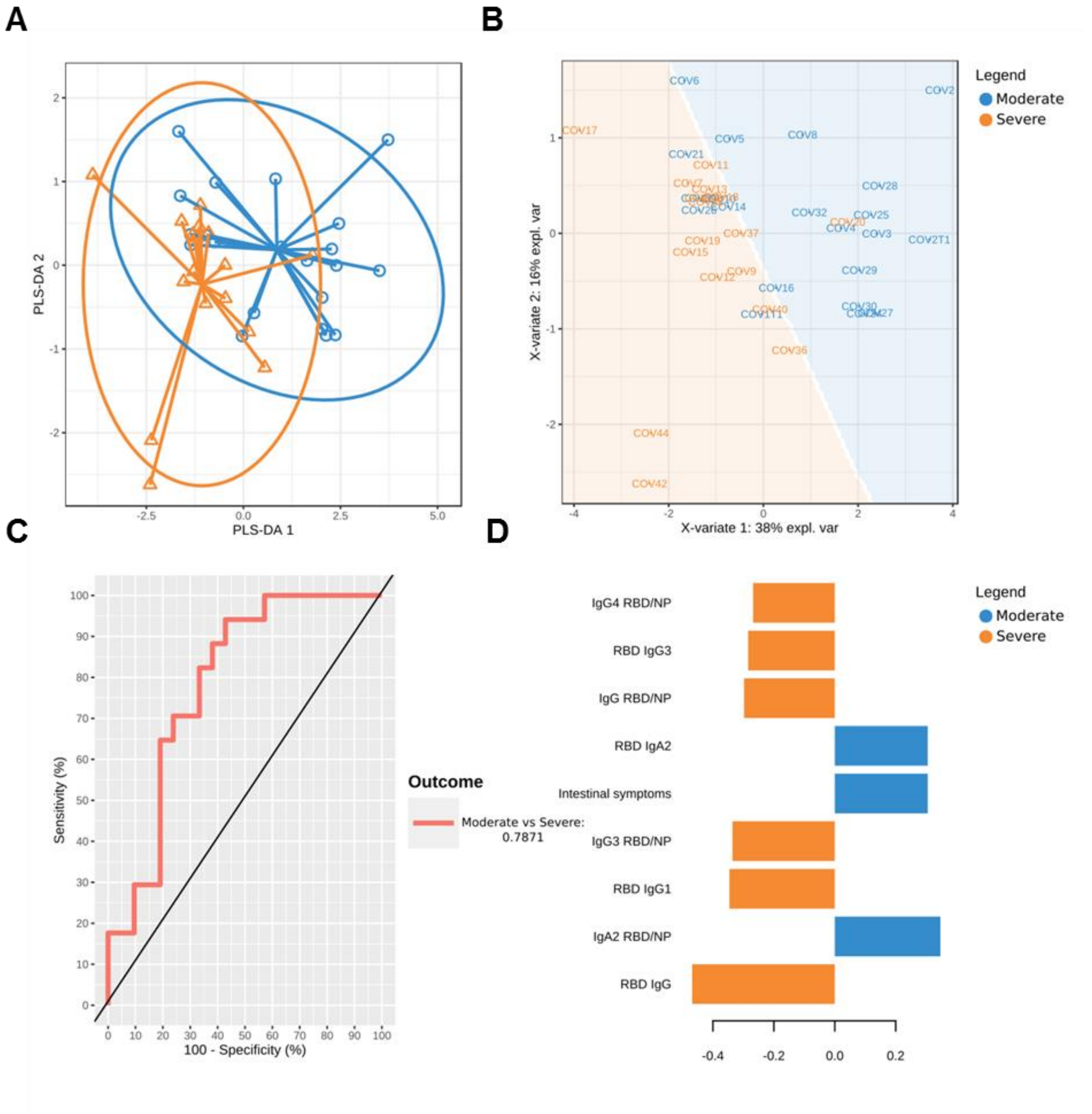


**Figure S6. Partial least squares-discriminant analysis (PLS-DA) of the antibody profiles and their ratios for COVID-19 patients.** The points are colored by sample group classification. (**A**) Map of the patients into the first two dimensions of the PLS-DA model. The points represent dimension scores of the projection of high-dimensional feature vectors onto the first (x axis) and second (y axis) dimensions. Confidence ellipses for each class are plotted to highlight the strength of the discrimination (confidence level set to 95%). (**B**) Map of the patients on the first two dimensions of the PLS-DA model with the 'prediction' background. The algorithm estimates the predicted area for each class, defined as the 2D surface where all points are predicted to be of the same class. (**C**) ROC Curve of the classification performance of the PLS-DA model for component one. The AUC is calculated from training cross-validation sets and averaged. (**D**) Loading weights of each variable on the first component of the PLS-DA model. The most important variables (according to the absolute value of their coefficients) are ordered from bottom to top.
